# Implementation of a Longitudinal Multimodal ECG Curriculum using Residents and Near-Peer Educators

**DOI:** 10.1101/2025.07.08.25330289

**Authors:** Tooba Alwani, Cancan Zhang, Kenneth J Mukamal, Gabriel P. Hurtado, Sakshi Wadhwa, Yoel Benarroch, Emily B. Crawford, Ian Kelly, Janet Monroe, Bence Zolyomi, Michael Gavin, Shu Yang, Jason Matos

## Abstract

**Introduction:** Electrocardiographic interpretation is an essential clinical competency but its proficiency is estimated at only 60% amongst internal medicine and emergency medicine residents. A recommended multimodal approach to electrocardiographic teaching includes longitudinal electrocardiographic exposure, near-peer teaching, vector-based interpretation, emphasis on common misinterpretations, clinical exposure, and novel teaching methods. Whether a longitudinal, asynchronous curriculum using residents as near-peer teachers can improve proficiency is uncertain.

**Research Question:** Does a resident-led, multimodal asynchronous electrocardiographic curriculum improve performance among internal medicine house-staff?

**Methods:** A total of 169 internal medicine house-staff at a single tertiary medical center were sent weekly emails containing a containing a preview electrocardiogram, video lesson, and relevant practice and learning points. Videos were created by second or third-year residents with cardiology faculty and posted on YouTube. To evaluate curriculum efficacy, we emailed residents a 14-item multiple-choice midterm assessment that tested respondents on electrocardiographic topics, including covered topics and not-yet-introduced material as controls. We compared the likelihood of correct responses to covered vs uncovered material in multivariable generalized estimating equation models with a logit link, adjusting for interest in cardiology and year of residency, with response correctness as the binary outcome for each item. We report odds ratios (ORs) and associated 95% confidence intervals (CIs).

**Results:** Video view counts declined over time, dropping from 171 to 19 instances from the first to last video. A total of 26 respondents (15.4%) completed the midterm assessment. Participants performed better on covered than uncovered topics (OR 1.66, 95% CI [1.07, 2.57]), but this association was similar among participants who did and did not report watching any videos (p interaction = 0.78).

**Conclusions:** A resident-led, multimodal ECG curriculum can be successfully developed, but low midterm assessment participation limited our ability to draw definitive conclusions about effectiveness. Declining engagement over time suggests that frequent email-based delivery may contribute to email fatigue and reduced participation. Future iterations should focus on enhancing engagement through alternative delivery methods, such as in-person or hybrid formats, and aim to increase participation to better assess effectiveness.

## Introduction/Background

Electrocardiographic (ECG) interpretation is a critical diagnostic tool for cardiovascular diseases such as ischemia, arrhythmias, and structural heart disorders^1^. Despite its importance, proficiency remains low, averaging 60% among internal and emergency medicine residents^2^ and 56% across various healthcare professionals^3^.

Although improving ECG education is vital, no universally accepted teaching method exists^4^. However, certain strategies show promise: lecture- and workshop-based learning outperform self-directed study^5^, summative assessments may enhance learning^6^, contrastive approaches aid pattern recognition^7^, and spaced repetition improves retention^8^.

A recent review advocates for a multimodal ECG curriculum incorporating longitudinal exposure, near-peer teaching, interactive learning, vector-based interpretation, clinical application, and novel techniques^9^. While some evidence supports this approach^9^, few studies have assessed its effectiveness—especially using medical residents as instructors. One study found that a fellow-led, longitudinal curriculum improved ECG interpretation and learner confidence among residents^10^.

This study aims to design and evaluate a longitudinal, multimodal ECG curriculum led by second- and third-year medical residents as near-peer instructors. The curriculum combines video lessons, asynchronous content, in-person sessions, and spaced repetition over several months to enhance ECG interpretation skills and comfort.

## Methods

### Sample

The study was reviewed and determined not to constitute human subjects research by the Committee on Clinical Investigations of Beth Israel Deaconess Medical Center. The sample included all 169 internal medicine house-staff, who received weekly curriculum emails.

### Curriculum

Second- and third-year residents with an interest in cardiology and medical education were invited via email to participate as content creators. Those interested selected one of the following topics to develop a video lesson: ECG basics, left/right ventricular hypertrophy (LVH/RVH) and strain, ischemia, atrioventricular nodal reentrant tachycardia (AVNRT)/ atrioventricular reentrant tachycardia (AVRT), atrial fibrillation and flutter, bradycardia, bundle branch blocks (BBB), or wide complex tachycardia (WCT). Each participating resident partnered with a faculty mentor to serve as an expert discussant in their lesson. These video lessons used a vector-based approach to explain key concepts and included several ECG examples for practice. The videos were generally 15-20 minutes in length.

The curriculum spanned eight months and followed a multimodal, longitudinal design delivered primarily via weekly emails. Each week, residents received a preview ECG, followed the next week by link to a video lesson created by resident teachers in collaboration with faculty. After each video, residents received a summary of key learning points along with an additional practice ECG that reinforced current and previous topics to support spaced repetition. Two in-person workshops were held midway through the curriculum to provide an interactive review of ECGs from previous sessions and occurred after the mid-term assessment (Figure 1).

**Figure 1:**
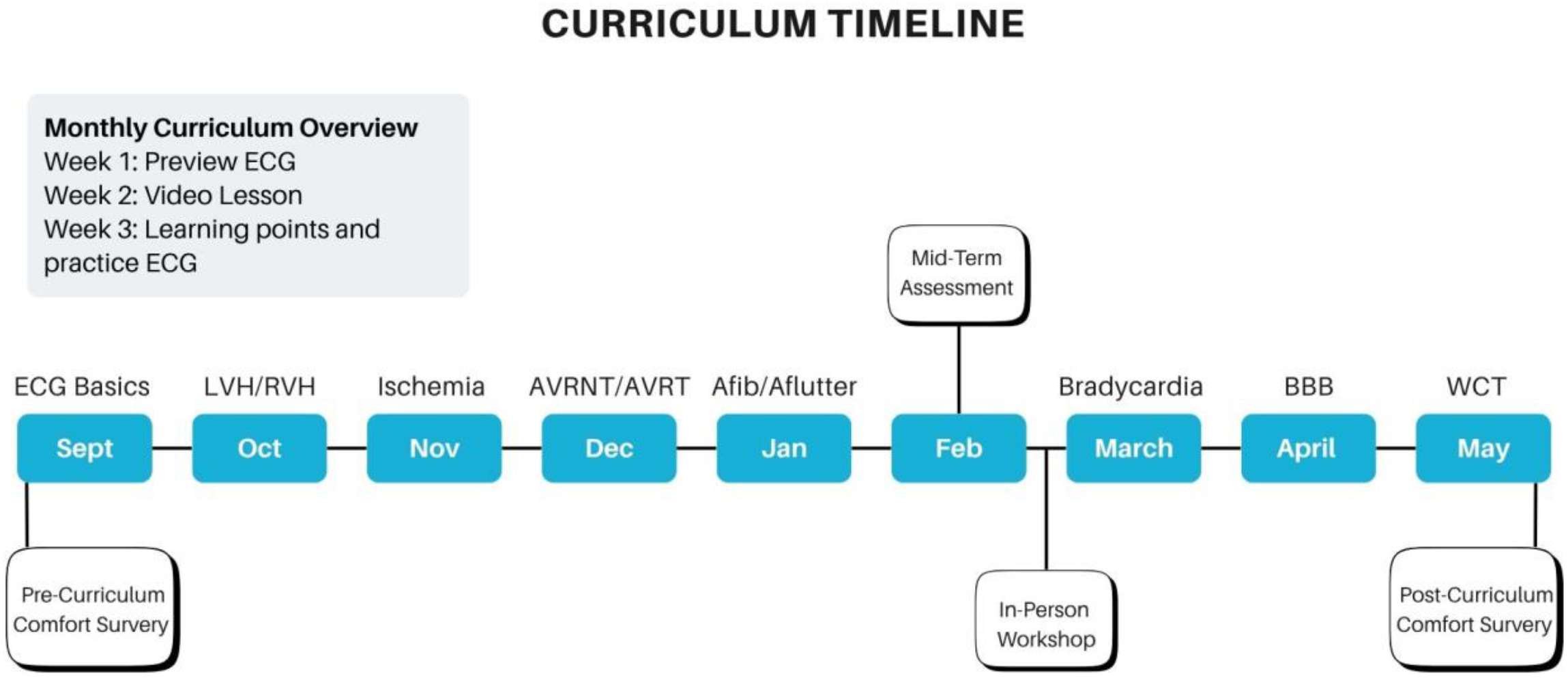
Curriculum Timeline. Shown here is a curriculum timeline highlighting the timing of administered surveys and assessments, as well as the monthly curriculum format. Abbreviations as follows: ECG (Electrocardiogram), LVH/RVH (Left Ventricular Hypertrophy/Right Ventricular hypertrophy, AVNRT (Atrioventricular Nodal Reentrant Tachycardia), AVNRT (Atrioventricular Reentrant Tachycardia), Afib (atrial fibrillation) and Aflutter (atrial flutter), BBB (Bundle branch blocks), WCT (Wide Complex Tachycardia).

In addition to the weekly emails, all materials were made accessible and remain so through a dedicated YouTube (channel (www.youtube.com/@Cardio-Learn), Google Drive, and later a dedicated website (https://www.cardio-learn.com/). Views were monitored based on reported public views on YouTube.

For purposes of evaluation, we randomized the order of video topics after the first lesson on ECG basics. Each lesson was presented as a stand-alone topic, mitigating any need for ordering.

### Assessment

To evaluate curriculum effectiveness, a 14-item multiple-choice midterm assessment was administered, covering seven ECG topics (with 2 questions per topic), including both introduced and not-yet-introduced material. As in previous assessments of educational material^11^, the uncovered material served as an internal control, and the primary measure of effectiveness was the likelihood of correct responses to covered versus uncovered material. Answers that were left blank were marked as incorrect.

Baseline and post-curriculum surveys using Likert-scale questions assessed resident comfort with ECG interpretation and with an anonymously created identifier to link pre- and post-survey responses. Given limited responses to the comfort survey, only descriptive statistics are presented.

### Statistical Analyses

We provide descriptive characteristics of the residents who completed the midterm assessment with counts and proportions. Additionally, we describe baseline characteristics with counts and proportions for residents who completed both the midterm assessment and the baseline survey. We assessed monotonic trends in the number of reported video views over time using the nonparametric Mann-Kendall trend test and report the associated p-value. To account for the multiple questions answered by each respondent, we used a multivariable generalized estimating equation (GEE) model with a logit link to analyze item-level correctness, adjusting for topic coverage, cardiology interest, and postgraduate year (PGY). We accounted for clustering within respondents using an exchangeable working correlation structure. Odds ratios (ORs) and 95% confidence intervals (CIs) were reported. We tested for an interaction between any video engagement and topic coverage using a multiplicative interaction term.

## Results

### Viewership

Online recorded views, as obtained by number of views on each YouTube video, ranged from 19 to 171 views, with decreasing views over time (Figure 2, Mann-Kendall trend test p=0.004). A total of 26 respondents (15.4%) completed the midterm assessment. Of the respondents, 17 (65 %) were not interested in pursuing cardiology, 2 (8%) were considering cardiology, and 7 (27%) were interested in cardiology. Regarding training level, 11 (44.0%) were PGY-1, 9 (36.0%) were PGY-2, and 5 (20.0%) were PGY-3. Of the respondents, 12 (46.2%) reported watching any of the available videos (Table 1).

**Table 1.** Descriptive characteristics of Mid-term Survey Respondents.

| Videos Watched # of people (percentage) |  |
| --- | --- |
| ECG basics | 11 (42.3%) |
| LVH/RVH | 6 (23.1%) |
| Ischemia | 4 (15.4 %) |
| AVNRT/AVRT | 3 (11.5%) |
| Afib/Aflutter | 3 (11.5%) |
| Any video | 12 (46.2%) |
| Interest in Cardiology |  |
| Yes | 7 (26.9% ) |
| No | 17(65.4%) |
| Maybe | 2 (7.7%) |
| PGY Year |  |
| 1 | 11 (44%) |
| 2 | 9 (36%) |
| 3 | 3 (5%) |

**LEGEND:** Shown are descriptive characteristics as well as reported videos watched by participants who responded to the mid-term assessment (n = 26). Data is reported as counts (percentage %). Abbreviations as follows: ECG (Electrocardiogram), LVH/RVH (Left Ventricular Hypertrophy/Right Ventricular hypertrophy, AVNRT (Atrioventricular Nodal Reentrant Tachycardia), AVNRT (Atrioventricular Reentrant Tachycardia), Afib (atrial fibrillation) and Aflutter (atrial flutter).

**Figure 2:**
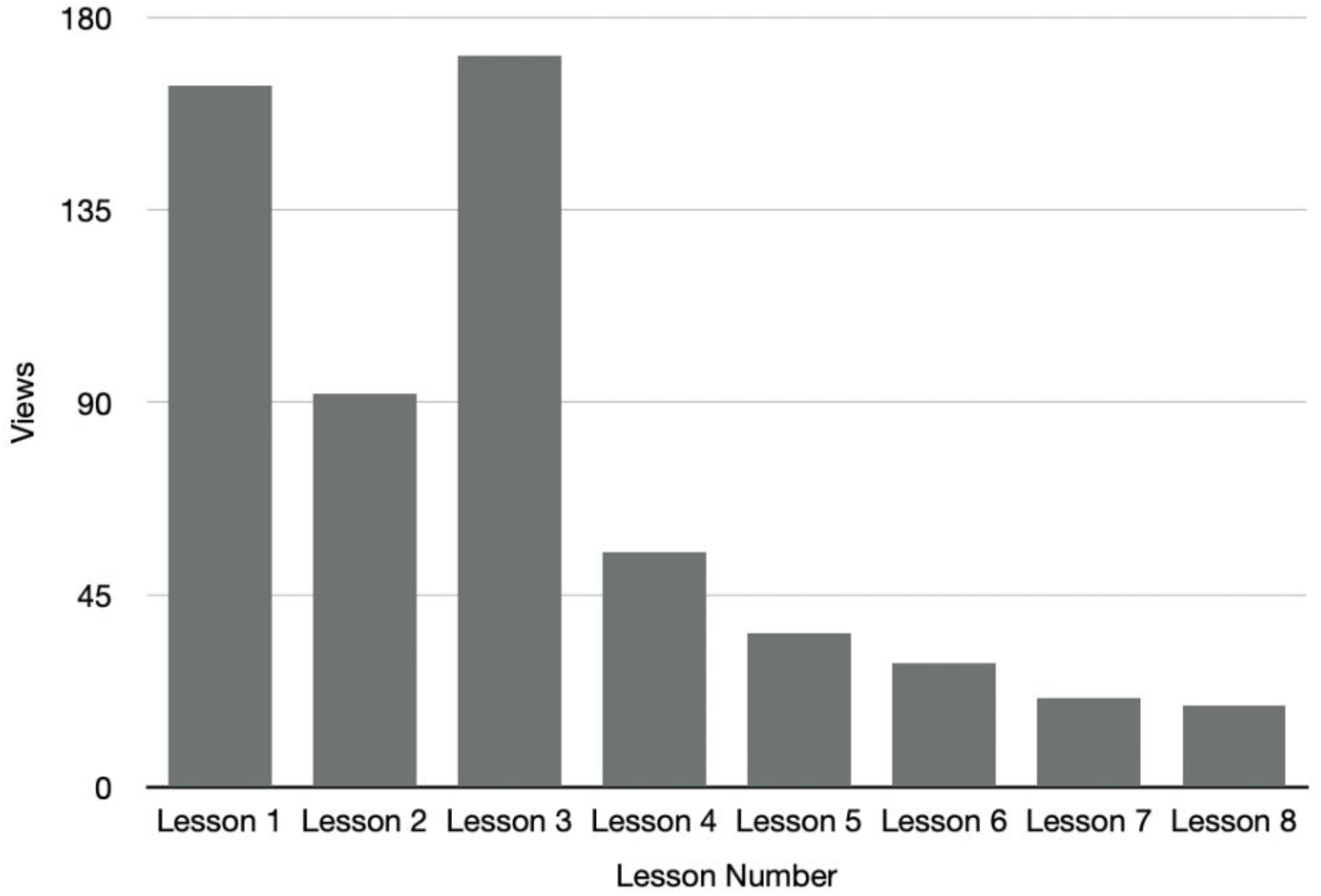
Video views over subsequent lessons. Shown here are the reported video views for each subsequent lesson (based on YouTube view counts) as the curriculum progressed throughout the year. The average number of views was 73.4 ± 62.7 (SD). Mann-Kendall trend test p=0.004.

### Performance

Overall, participants answered an average of 70% of questions correctly. For topics covered, they answered an average of 75% of the questions correctly, in comparison to 63% on uncovered topics (OR 1.66, 95% CI [1.07, 2.57]. To evaluate whether this perceived benefit of the curriculum might reflect differences in underlying difficulty of individual questions, we tested whether the effect of coverage differed among individuals who reported watching at least one video. We found no significant difference between groups (p multiplicative interaction 0.78), suggesting that viewing did not improve the likelihood of correct responses to covered vs uncovered material. We also observed no interaction by PGY year or interest in cardiology.

### Comfort

A total of 63 participants completed the baseline survey assessing comfort. Of these, 13 completed the mid-term assessment and are described below (Table 2).

**Table 2.** Descriptive characteristics of initial comfort survey respondents.

| Likert Scale Responses- Mean (SD) |  |
| --- | --- |
| LVH | 2.00 (0.91) |
| RVH | 1.77 (0.93) |
| Ischemia | 2.62 (1.12) |
| AVNRT/AVRT | 1.54 (0.66) |
| Afib/Aflutter | 3.23 (1.17) |
| Bradycardia | 4.08 (0.76) |
| Bundle branch blocks | 2.23 (1.42) |
| Wide complex Tachycardia | 3.85 (0.90) |
| Interest in Cardiology- Number (proportion) |  |
| Yes | 1 (8.3%) |
| No | 8 (66.7%) |
| Maybe | 3 (25%) |
| PGY Year- Number (proportion) |  |
| 1 | 7 (53.8) |
| 2 | 6 (46.2%) |
| 3 | 0 |

**LEGEND:** Shown are mean Likert scale responses for participants who responded to initial comfort survey and completed the midterm assessment (n = 13). Likert scale ranges from 1= very uncomfortable to 5 = very comfortable. Responses are reported as Mean (SD). Additionally descriptive characteristics are reported as counts (percentage %). Abbreviations as follows: ECG (Electrocardiogram), LVH/RVH (Left Ventricular Hypertrophy/Right Ventricular hypertrophy, AVNRT (Atrioventricular Nodal Reentrant Tachycardia), AVNRT (Atrioventricular Reentrant Tachycardia), Afib (atrial fibrillation) and Aflutter (atrial flutter).

## Discussion

This study demonstrates the feasibility of creating a resident-led, multimodal ECG curriculum within an internal medicine residency program. The curriculum incorporated evidence-based strategies such as near-peer teaching, spaced repetition, vector-based instruction, and a longitudinal structure. At the same time, the experience also demonstrates the challenges of implementation. While participants performed better on topics that had been covered, and the choice of topics was randomly ordered, the overall impact on ECG interpretation skills is difficult to quantify due to limited participation and declining engagement over time, and our results are consistent with the possibility that covered topics were, by chance, easier to answer correctly.

Although the curriculum showed potential in supporting learning—particularly through improved performance on covered topics—its evaluation was limited by a low number of assessment respondents (27 out of 169) and a steady decline in video engagement. While initial videos garnered many views, the number of completed assessments was disproportionately low, highlighting a critical gap between content consumption and measurable outcomes. This discrepancy underscores a key challenge in evaluating medical education curricula: passive engagement does not necessarily translate to active learning or reliable assessment.

The decline in engagement over time also reflects common challenges in asynchronous learning, including resident fatigue and reduced motivation as the academic year progresses. This is in line with prior studies which have shown that self-paced, video-based learning has a strong initial uptake followed by steep attrition as learners progress through modules without structured reinforcement^12^. Additionally, frequent email-based delivery may contribute to email fatigue, reducing sustained participation. A multi-institutional randomized study of an ECG curriculum delivered via email found that while novice learners benefited, overall knowledge gains were limited and participation declined as the modules progressed^13^. Another study showed demonstrated that spaced education emails improved retention among interns, but not among more senior residents—highlighting how engagement with email-based content can vary by level of training^11^. Together, these findings underscore how asynchronous formats, while convenient, may be insufficient on their own to maintain long-term engagement or ensure consistent learning outcomes without integrated accountability structures, interactive components, or faculty oversight.

Despite these limitations, the flexibility of a partially asynchronous curriculum remains essential given residents’ demanding schedules. Future iterations should explore hybrid models—such as integrating content into academic half-days, offering periodic in-person workshops, or using app-based platforms—to enhance both engagement and accessibility. Efforts should also focus on increasing response rates through interactive activities or financial incentives and incorporating more robust outcome measures, such as summative assessments delivered during mandatory didactic sessions, to enable more accurate and meaningful evaluation.

In summary, while a resident-led, multimodal ECG curriculum is both feasible to develop and well-aligned with best practices in medical education, further refinement is needed to enhance implementation and engagement and assess effectiveness on a larger scale.

## Data Availability

All data produced in the present study are available upon reasonable request to the authors

## References

1. Rafie N, Kashou AH, Noseworthy PA. ECG Interpretation: Clinical Relevance, Challenges, and Advances. Hearts. 2021;2(4):505–513. doi:10.3390/hearts2040039

2. Berger JS, Eisen L, Nozad V, et al. Competency in electrocardiogram interpretation among internal medicine and emergency medicine residents. Am J Med. 2005;118(8):873–880. doi:10.1016/j.amjmed.2004.12.004

3. Kashou AH, Noseworthy PA, Beckman TJ, et al. ECG Interpretation Proficiency of Healthcare Professionals. Curr Probl Cardiol. 2023;48(10):101924. doi:10.1016/j.cpcardiol.2023.101924

4. Fent G, Gosai J, Purva M. Teaching the interpretation of electrocardiograms: Which method is best? J Electrocardiol. 2015;48(2):190–193. doi:10.1016/j.jelectrocard.2014.12.014

5. Mahler SA, Wolcott CJ, Swoboda TK, Wang H, Arnold TC. Techniques for teaching electrocardiogram interpretation: self-directed learning is less effective than a workshop or lecture. Med Educ. 2011;45(4):347–353. doi:10.1111/j.1365-2923.2010.03891.x

6. Raupach T, Brown J, Anders S, Hasenfuss G, Harendza S. Summative assessments are more powerful drivers of student learning than resource intensive teaching formats. BMC Med. 2013;11(1):61. doi:10.1186/1741-7015-11-61

7. Hatala RM, Brooks LR, Norman GR. Practice makes perfect: the critical role of mixed practice in the acquisition of ECG interpretation skills. Adv Health Sci Educ Theory Pract. 2003;8(1):17–26. doi:10.1023/a:1022687404380

8. Jm C, M J, T K, et al. The spacing effect: Improving electrocardiogram interpretation. Clin Teach. 2024;21(1). doi:10.1111/tct.13626

9. Kaye MG, Kwiatkowski AV, Khan HA, Yastynovich Y, Graham SP, Meka J. Designing an ECG curriculum for residents: Evidence-based approaches to improving resident ECG interpretation skills. J Electrocardiol. 2024;82:64–68. doi:10.1016/j.jelectrocard.2023.10.012

10. Kaye MG, Khan HA, Gudleski GD, Yatsynovich Y, Graham SP, Kwiatkowski AV. Implementation of a longitudinal, near-peer ECG didactic curriculum in an internal medicine residency program and impact on ECG interpretation skills. BMC Med Educ. 2023;23(1):526. doi:10.1186/s12909-023-04483-y

11. Matos J, Petri CR, Mukamal KJ, Vanka A. Spaced education in medical residents: An electronic intervention to improve competency and retention of medical knowledge. PloS One. 2017;12(7):e0181418. doi:10.1371/journal.pone.0181418

12. Fernandez A, Moeller JJ, Harrar DB, et al. Curriculum Innovation: Design and Implementation of Synchronous and Asynchronous Curricula to Enhance Residents’ EEG Knowledge and Experience. Neurol Educ. 2023;2(4):e200101. doi:10.1212/NE9.0000000000200101

13. Klein AJ, Berlacher M, Doran JA, Corbelli J, Rothenberger SD, Berlacher K. A Resident-Authored, Case-Based Electrocardiogram Email Curriculum for Internal Medicine Residents. MedEdPORTAL. 16:10927. doi:10.15766/mep_2374-8265.10927

